# Prediction of severe COVID-19 cases requiring a ventilator in Tokyo, Japan

**DOI:** 10.1101/2021.03.11.21253431

**Authors:** Junko Kurita, Tamie Sugawara, Yasushi Ohkusa

**Author notes:** **Corresponding author:** Junko Kurita.

## Abstract

**Background:** To avoid exhaustion of medical resources by COVID-19, policy-makers must predict care needs, specifically the proportion of severe cases likely to need ventilator care.

**Objective:** This study was designed to use a statistical model to elucidate dynamics of severe cases in Tokyo and to discuss the timing of effective policy activation.

**Methods:** The study extended from April 27 through October 18, 2020 in Japan’s Tokyo Metropolitan area. Medical exhaustion was defined as use of more than half of the ventilator capacity available before the COVID-19 outbreak. We regressed the number of severe cases on the newly onset patients of more than 14 days prior. As earlier research indicated, the COVID-19 severity changed at the end of May. Therefore, we added dummy variables to reflect changing severity from June and its product with newly onset patients as the explanatory variable. Then we calculated the threshold using R(*t*): R(*t*)=0.99 for the number of patients 14 days prior was used as a threshold at which strong countermeasures should be activated to keep to avoid medical exhaustion.

**Results:** The critical number signaling medical exhaustion in Tokyo was defined as 655 cases. We selected 15, 30, 60 and 90 days prior as explanatory variables for explaining the number of severe cases. A coefficient of determination larger than 0.95 was inferred as indicating good fit. The threshold was estimated as more than 4500 cases for R(*t*)=1.1 and monotonically decreasing by R(*t*) to be 600 cases for R(*t*)=2.5.

**Discussion and Conclusion:** Results showed that newly onset patients reported more than 14 days prior can explain the number of severe cases very well. We also confirmed the threshold number of patients 14 days prior by R(*t*) for which strong countermeasures should be activated to avoid medical exhaustion with R(*t*)=0.99. This method is expected to be useful for countermeasure activation policies for Tokyo.

## Introduction

As of October 28, 2020, the World Health Organization reported more than 42 million confirmed COVID-19 cases and about 1.15 million deaths worldwide (1). In Japan, the outbreak reached its peak at the end of July. By October 29, 2020, approximately 100 thousand cases and 1.7 thousand deaths had been reported (1).

For the 29,601 cases reported from February 1 through August 5, 2020, the case–fatality rate (CFR) was low for people 59 years old and younger. Nevertheless, among patients 60 years old and older, CFR was found to increase with age. Similarly, probabilities of ICU admission and ventilator use increased rapidly with age (2). Patients with underlying diseases such as hypertension or diabetes constituted high-risk groups for severe cases of COVID-19 (3,4).

The Japanese government set guidelines for countermeasures against COVID-19 on August 28, emphasizing caution for elderly people and high-risk individuals with underlying diseases (5). Based on those guidelines, medical resource allocations thereafter emphasized high-risk groups, including elderly people, rather than patients with mild symptoms or asymptomatic patients.

Given these circumstances, understanding and predicting the proportion of severe cases requiring ventilator treatment are necessary for policy makers if a strong policy requiring self-restriction of travel is to be made, or if customer-service businesses must be closed. Therefore, this study was conducted to elucidate the dynamics of severe cases in Tokyo, with discussion of the timing of activating countermeasure policies.

Although we applied mathematical models to outbreaks of COVID-19 in Japan, such models are unable to explain why the outbreak reached a peak at the end of July under conditions of no herd immunity or lock-down. Actually, at that time in late July, the Japanese government had encouraged domestic travel and thereby promoted the spread of the outbreak. Nevertheless, the outbreak was contained in August. Therefore, to date, no mathematical model has been able to explain the outbreak in Japan. Instead, we examined a statistical model rather than a mathematical model to explain the course of the outbreak. We propose some thresholds that are useful to signal activation of strong countermeasures.

## Methods

Severe cases of COVID-19 were defined as requiring hospitalization of patients with required use of a ventilator. The study period extended from April 27, when severe cases under the current definition began to be reported in Tokyo, to October 18, 2020. The Tokyo Metropolitan area of Japan was the study area. Medical exhaustion was defined as patients using more than half of the ventilator capacity available before the COVID-19 outbreak.

We regressed the number of severe cases on the newly onset patients reported more than 14 days prior. As earlier studies have indicated, the severity of COVID-19 changed at the end of May. Therefore, we added dummy variables to reflect changing severity from June and its product with newly onset patients as an explanatory variable. We selected a formula maximizing the adjusted coefficient of determination.

Then, we calculated the threshold by assuming constant R(*t*) after October 19, 2020. The timing of R(*t*)=0.99 was assessed for activation of strong countermeasures to avoid medical facility exhaustion, with activation occurring when the number of patients 14 days prior reached this criterion level. Medical facility exhaustion was defined as patients requiring ventilator use exceeding half of the ventilator capacity available before the COVID-19 outbreak.

The study period for the epidemic curve of symptomatic patients was January 16, 2020 through October 18, 2020. For severe cases, it was April 27, 2020, when severe cases were defined as patients who needed ventilators, through October 18, 2020. Because incubation periods and delays of reporting varied after onset of symptoms, data of the prior month were expected to be underreported. Therefore, we did not use the final month of data available to us. However, collection of data of patients using a ventilator started April 27 in Tokyo. Therefore, data of ventilator usage on or before April 26 were not used for this study. The study area was metropolitan Tokyo, with 14 million residents. The epidemic curve and information about severe cases in Tokyo were provided by the Tokyo Metropolitan Government (6).

We also performed out of sample prediction assuming that R(*t*) will be constant from October 19, 2020. We calculated the threshold to activate strong policy measures for reducing R(*t*) to be 0.99 for the number of newly onset patients in a day in terms of R(*t*). Such measures might include some requirements of voluntary restrictions against going out of the home or shutting down customer-service businesses. Under such policies, one can expect to avoid exhaustion of medical resources and keep the need for ventilators lower than 655.

Finally, to evaluate the model’s predictive power, we performed out of sample forecasting prospectively for 21 days from December 3, 2020. The parameters for forecasting were fixed. The estimation result used data obtained through October 18, 2020.

### Ethical considerations

All information used for this study was collected under the Law of Infection Control, Japan. Published data were used. Therefore no ethical issue is posed by the procedures used to conduct this study.

## Results

In Tokyo, we observed 3,072 patient days of ventilator use from April 27, 2020 through the end of July. From January 16, 2020 through the end of July, 11,619 patients with symptoms were reported.

Because Tokyo had 2,456 ventilators before the COVID-19 outbreak and because 1,310 ventilators were unused by patients (7), if patients needing ventilators were to become more numerous than 655, the situation would be designated as medical facility exhaustion.

Figure 1 depicts severe cases and newly onset patients. Severe cases fluctuate with a longer lag than the onset patients. The estimation results are presented in Table 1. Because 120 days before April 27 was earlier than January 16, when the initial case in Tokyo showed onset, the specification with onset patients 120 days prior includes fewer observations than other specifications. Based on adjusted coefficients of determination, we selected the model with up to 90 days prior. Its adjusted coefficient of determination was higher than 0.95. Figure 2 depicts the observed and fitted numbers of severe cases. It is apparent that these were almost equal to one another.

**Table 1:** Estimation result for the number of sever cases

| Explanatory variable | Estimated coefficients | *p*-value | Estimated coefficients | *p*-value | Estimated coefficients | *p*-value | Estimated coefficients | *p*-value |
| --- | --- | --- | --- | --- | --- | --- | --- | --- |
| constant | 3.899 | 0.004 | 3.821 | 0.025 | 5.743 | 0.000 | 11.549 | 0.000 |
| t-15 | 0.011 | 0.206 | 0.024 | 0.002 | 0.021 | 0.017 | -0.007 | 0.377 |
| t-30 | 0.064 | 0.000 | 0.059 | 0.000 | 0.054 | 0.000 | 0.073 | 0.000 |
| t-60 | 0.030 | 0.000 | 0.029 | 0.000 | 0.031 | 0.000 |  |  |
| t-90 | 0.013 | 0.020 | 0.012 | 0.084 |  |  |  |  |
| t-120 | 0.020 | 0.079 |  |  |  |  |  |  |
| dummy variable for after June | 19.256 | 0.000 | 15.617 | 0.000 | 1.272 | 0.625 | 3.097 | 0.083 |
| Cross term with dummy variable for after June |  |  |  |  |  |  |  |  |
| t-15 | 0.059 | 0.545 | 0.302 | 0.000 | 0.229 | 0.000 | 0.175 | 0.000 |
| t-30 | 0.268 | 0.000 | 0.218 | 0.000 | 0.329 | 0.000 | 0.300 | 0.000 |
| t-60 | -0.047 | 0.008 | -0.031 | 0.041 | 0.030 | 0.058 |  |  |
| t-90 | -0.101 | 0.000 | -0.135 | 0.000 |  |  |  |  |
| t-120 | -0.477 | 0.240 |  |  |  |  |  |  |
| Number of observations = | 157 |  | 175 |  | 175 |  | 175 |  |
| Adjusted coefficients of determinant | 0.8829 |  | 0.9613 |  | 0.9443 |  | 0.9265 |  |

Note: t-x (x=15,30,60,90, and 120) indicates the number of newly onset patients x days before. Panel lower than “Cross term with dummy variable for after June” shows cross term with dummy variable for after June and he number of newly onset patients x days before. The most left specification with t-120 has fewer observations than other specification because 120 day before from April 27 was earlier than January 16 when the initial case in Tokyo was onset. Based on adjusted coefficients of determinant, we selected the model with up to 90 days before

**Figure 1:**
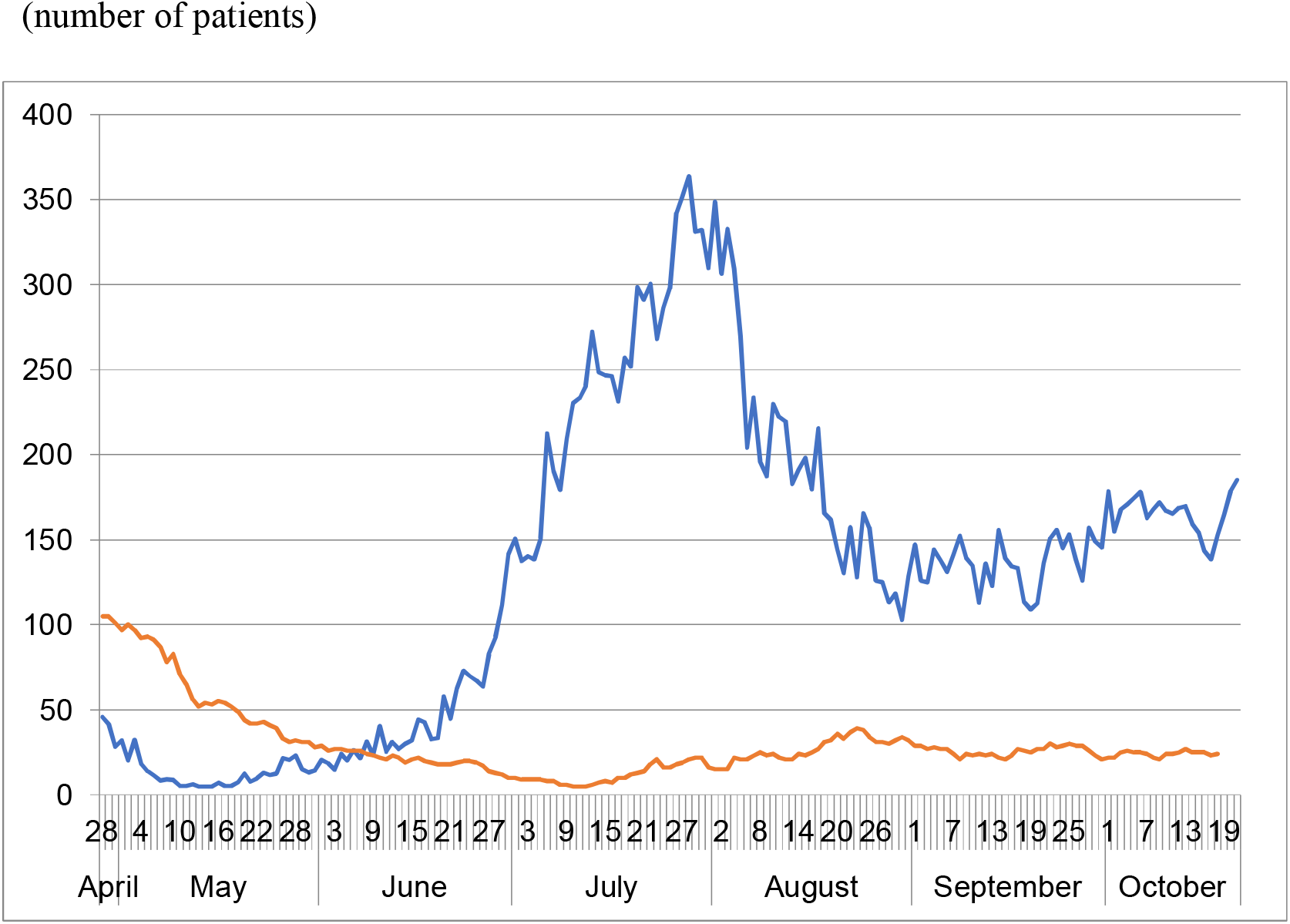
Epidemic curve of the symptomatic confirmed cases and number of patients requiring a ventilator in Tokyo, April 28 – October 18, 2020. Notes: The blue line shows the epidemic curve of the symptomatic confirmed cases on the onset date. The red line represents the number of patients with a ventilator.

**Figure 2:**
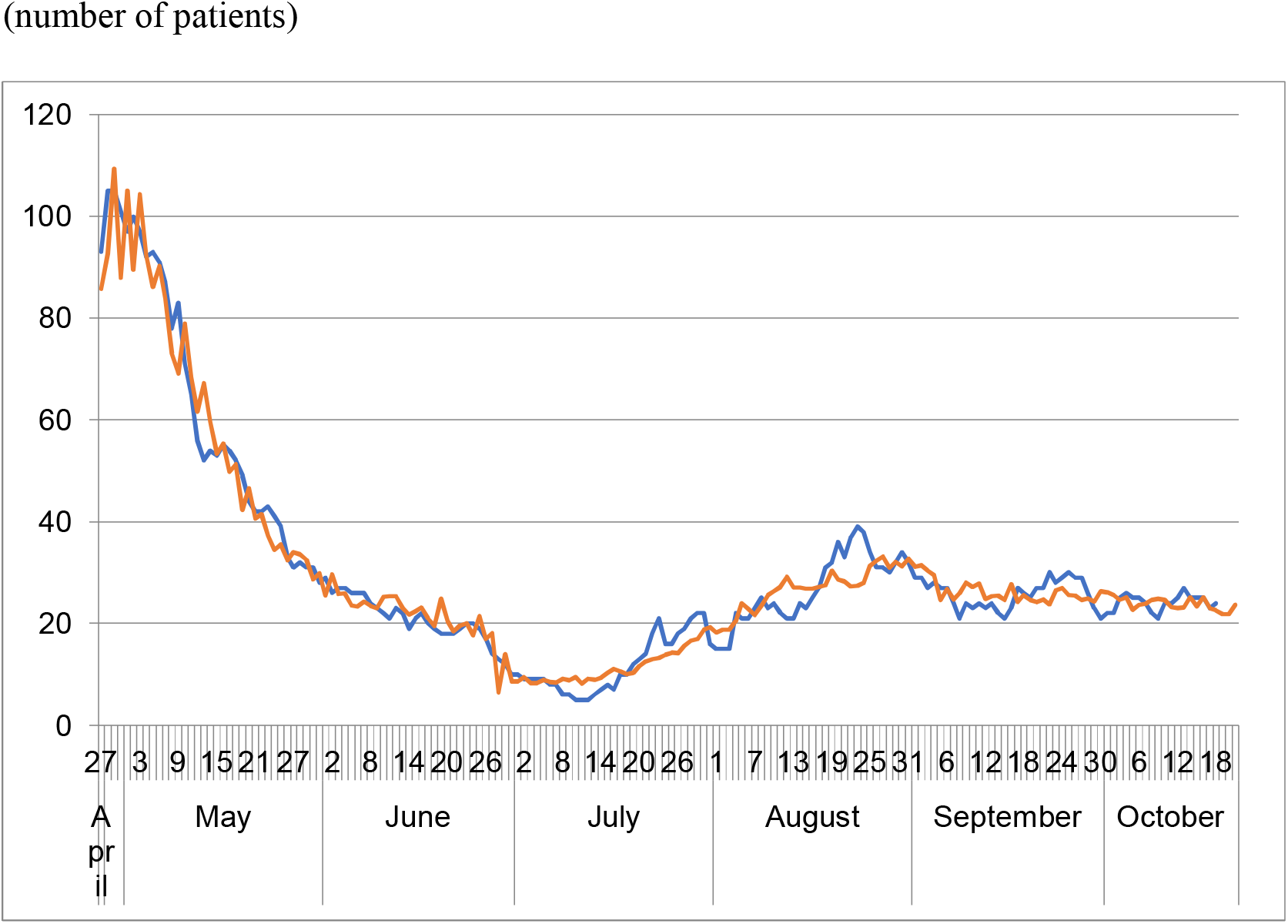
Estimated and observed number of patients requiring a ventilator in Tokyo, April 28 – October 18, 2020. Note: Blue The blue line shows the observed number of patients requiring a ventilator. The red line represents the estimated number of patients requiring a ventilator.

The estimated thresholds of the number of patients 14 days prior by R(*t*) in which the timing of activation of strong countermeasures to be R(*t*)=0.99 should be activated to avoid medical exhaustion are shown at Figure 3. The threshold was estimated as more than 4500 for R(*t*)=1.1, decreasing monotonically by R(*t*) to be 600 for R(*t*)=2.5.

**Figure 3:**
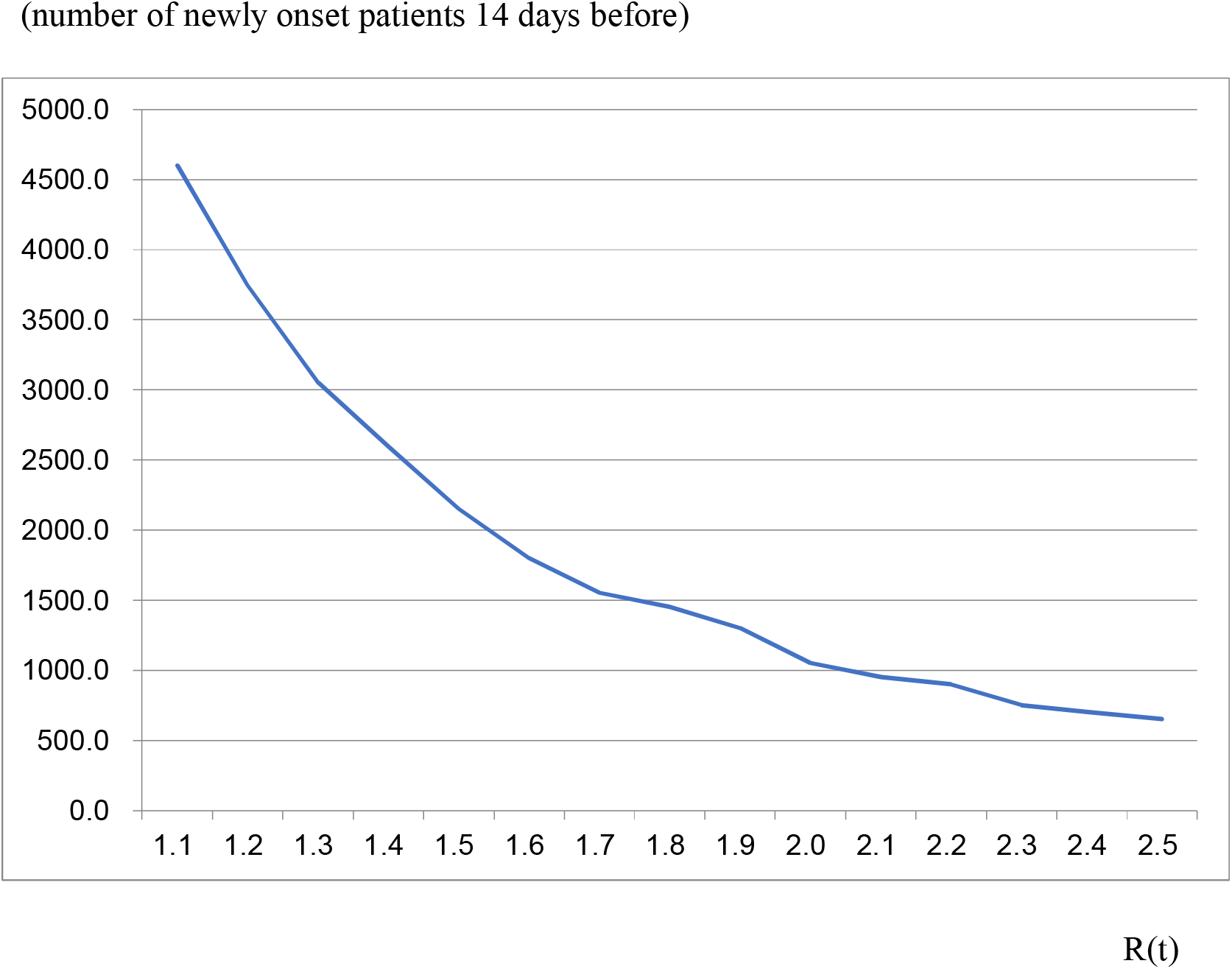
Estimated threshold of the number of newly onset cases 14 days prior to avoid medical facility exhaustion by R(*t*). Notes: The line represents the threshold of the number of newly onset patients on 14 days prior, by R(*t*). Medical facility exhaustion will occur if newly onset patients during 14 days prior exceed the threshold.

Figure 4 depicts out of sample forecasting prospectively for 21 days from December 3, 2020 through February 23, 2021. The observed peak was January 20, 2021, but the peak in forecasting was January 29, 2021. Before the peak date, forecasting was significantly lower than the observed figure, except for early December. However, after the peak date, no significant difference was found among them.

**Figure 4:**
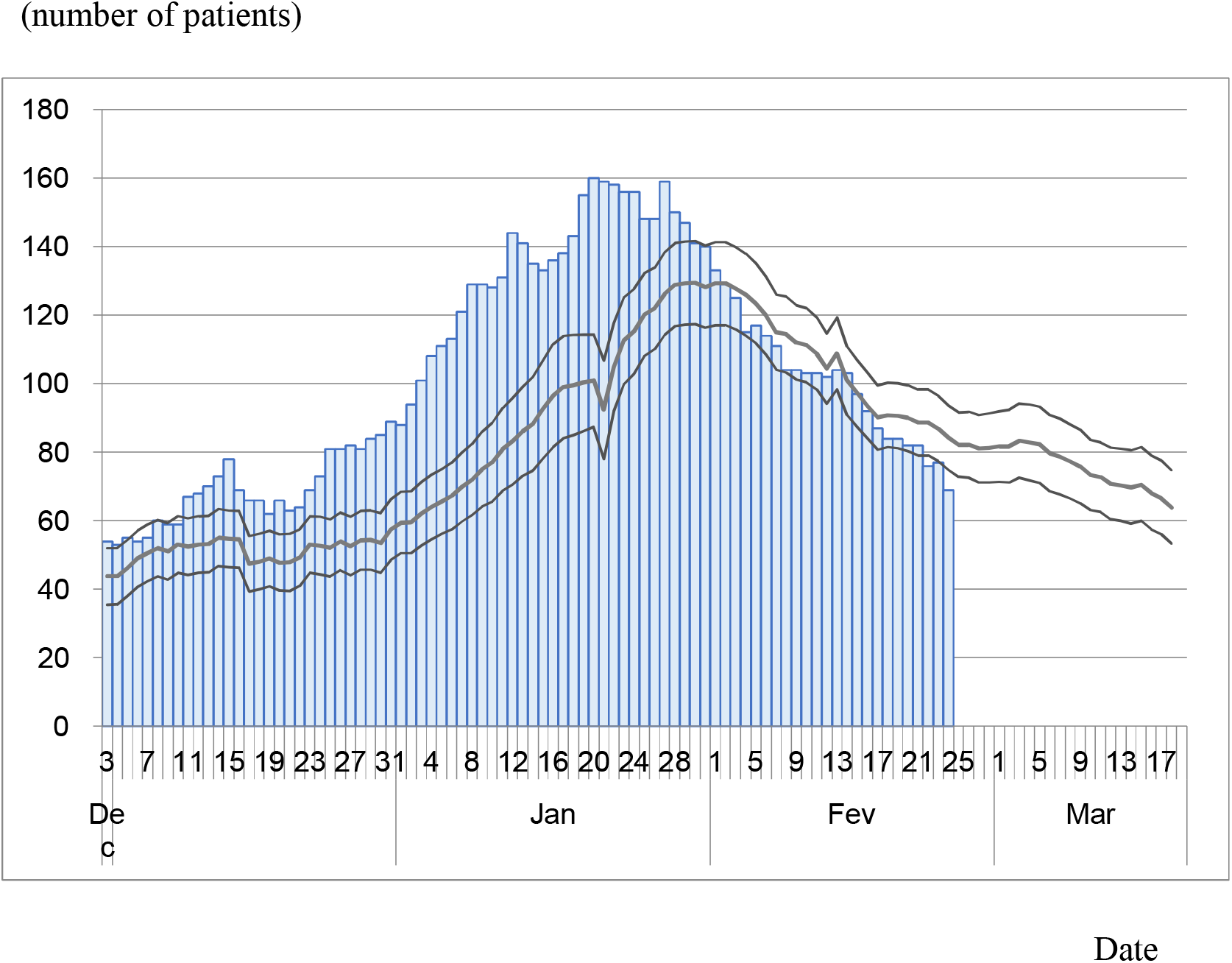
Observed data of severe cases and its forecasting. Notes: Bar represents data of severe cases observed in Tokyo from December 3, 2020. The bold line represents a forecast based the estimation results presented in Table 1. Thin lines represent its 95% CI.

## Discussion

Many mathematical models have been applied to the outbreak of COVID-19 worldwide (8,9). Unfortunately, these have not succeeded in explaining the course of COVID-19 outbreaks.

Especially, in Japan, two peaks were experienced through October. The first peak was that of earlier April; the second one was at the end of July. The first peak might explain the mathematical model with restriction of going out (10) because, since mid-March, the rate of going out declined, as shown in Apple data. However, the second peak is inexplicable by the mathematical model. Herd immunity, of course, was not achieved. No lock-down was activated in Japan at all. Probably, the “new normal” lifestyle with mask-wearing, social distancing, and restrictions against numerous dining or night-life establishments became common.

Actually, at that time, the government encouraged domestic travel. It promoted the spread of the outbreak. However, the outbreak was contained in August. Therefore, to date, no mathematical model has been sufficient to explain the outbreak in Japan. The policy for severe cases cannot be discussed using mathematical models.

Instead, we used a statistical model. It might always show some association of variables for which there is no causality. We showed clear association among severe cases and patients who had experienced onset 15 days prior. Moreover, we showed the threshold of the number of patients 14 days prior by R(*t*), for which the timing of activation of strong countermeasures to be R(*t*)=0.99 should be activated to avoid medical exhaustion. Therefore, the statistical model was probably more useful than the mathematical model for countermeasures against COVID-19.

Regarding the increase phase in June and July, the number of newly onset patients increased from about 10 to 350 during 60 days. If one assumes an average incubation period to be about 6 (10), then the average R(*t*) during this period was 1.43=(350/10)^(6/60). Figure 3 depicts the threshold as around 2200 at R(*t*)=1.43. The peak at the end of July was 350. Therefore, the peak at the end of July accounted for only 16% of the threshold.

We used no information before *t*-14. Reporting was usually delayed from onset up to 31 days (10). Therefore, the number of newly onset patients at the same day was that which was usually developing dynamically for at least two weeks. Information of the final two weeks prior to the estimate is not reliable. Therefore, we ignored information during the final two weeks of the study period. Moreover, because the epidemic curve changes gradually, information related to day *t* is closely correlated with information related to day *t*-1. Therefore, we used no information related to consecutive days. We used the number of newly onset patients with 15–30 day intervals as explanatory variables.

Prospective out of sample forecasting in Figure 4 shows good fit. Especially in the declining phase, it fits very well. Therefore, results demonstrated that the estimated model can predict outcomes well for 21 days.

Some limitations might have affected this study. First, we used dummy variables after June to reflect the disease’s decreasing severity. However, we do not understand the reason why. It might be attributable to mutations of the COVID-19 virus. Alternatively, treatment for patients might have improved since June. Or younger and therefore milder patients accounted for a larger proportion of the patients than before June. Moreover, the severity of the disease would decrease if a new drug were developed. It might alternatively increase if some mutation were to strengthen its pathogenicity. Severity should be monitored to improve the fit of the statistical model.

Secondly, definitions of severe cases differed among prefectures, even in Japan. Moreover, they probably varied among countries. It remains unknown whether the approach used for the present study is applicable to other prefectures than Tokyo or other countries with other definitions of severity. We should check the robustness of usefulness of the statistical model in other areas in Japan or other countries.

Thirdly, we might specifically examine only elderly patients or patients older than 50 as explanatory variables. If the reason for lower severity after June was the larger proportion of younger patients, then it might be controlled by specifically examining elderly patients as a high risk group. Exploration of that point remains as a challenge for future research efforts.

## Conclusion

Results demonstrated that newly onset patients more than 14 days prior can explain the number of severe cases very well. The prospective out of sample forecasting used for this study can predict observations well for 21 days. Results also show a threshold of the number of patients 14 days prior by R(*t*). Timing of the activation of strong countermeasures should be R(*t*)=0.99 to avoid medical facility exhaustion. That signal might be useful for countermeasure activation in Tokyo. Careful real-time monitoring of R(*t*) and newly onset patients are expected to be extremely important for timing of the activation of strong policy measures.

The present study is based on the authors’ opinions. Its results do not reflect any stance or policy of their professionally affiliated bodies.

## Data Availability

WHO Coronavirus Disease (COVID-19) Dashboard 
Cabinet Secretariat. COVID-19 Information and Resources.(in Japanese)

https://covid19.who.int/

https://corona.go.jp/dashboard.

## Acknowledgments

We acknowledge the great efforts of all staff at public health centers, medical institutions, and other facilities who are fighting the spread and destruction associated with COVID-19. This study represents the authors’ opinion. It does not reflect any stance of our affiliation.

